# Virus testing optimisation using Hadamard pooling

**DOI:** 10.1101/2024.10.21.24315883

**Authors:** Godfrey S. Beddard, Briony A. Yorke

## Abstract

Pooled testing is an established strategy for efficient surveillance testing of infectious diseases with low prevalence. Pooled testing works by combining clinical samples from multiple individuals into one test, where a negative result indicates the whole pool is disease free and a positive result indicates that individual testing is needed. Here we present a straightforward and simple method for pooled testing that uses the properties of Hadamard matrices to design optimal pooling strategies. We show that this method can be used to efficiently identify positive specimens in large sample sizes by simple pattern matching, without the requirement of complex algorithms.

## Background

Many countries have used the ‘test, trace and isolate’ strategy to control the spread of highly infectious diseases such as SARS-CoV-2. The global capacity for clinical testing is limited by factors including cost, availability of reagents and testing capacity of clinical laboratories. The gold-standard for detecting SARS-CoV-2 is RT-PCR [1], this technique is expensive, requires specialized equipment and reagents, and is relatively slow, taking a few hours per test on average [2, 3].

During the height of the COVID-19 pandemic lateral flow testing (LFT) provided a solution to issues regarding laboratory testing capacity but were not without limitations. The ‘test, trace and isolate strategy’ relies on accurate, reproducible testing and data monitoring, however, LFT self-testing is hindered by test variability, false negatives, and data loss due to errors in self-reporting [4]. In combination, SARS-CoV-2 testing resulted in tens of thousands of tons of plastic waste [5], an unsustainable consumption of reagents [6] and gold nanoparticles [7].

Strategies to address this issue have resulted in the development of biodegradable and recyclable biosensors [8, 9], methods to recover gold nanoparticles [10] and microfluidic and reagent-free testing devices [11–14]. Despite these developments, individual testing remains inefficient when the fraction of positives in a population is small, i.e. when almost all tests are expected to be negative. However, this is just the situation that needs surveillance-type testing, so that a new pandemic can be anticipated. When the infection rate is lower and urgency is not so pressing, one way of increasing capacity is to group or pool samples. This batch analysis was first suggested by Dorfman [15] more than 70 years ago, but has received renewed interest due to the critical testing bottlenecks that were highlighted during the COVID-19 pandemic [16–18].

### Sample Pooling

Dorfman’s method consists of two stages, the first is to combine samples from multiple individuals for analysis. If this test result is negative then all samples are negative, if the test is positive for the pool then each sample is tested again. The efficiency of this method depends on the positivity rate and can be improved by applying various strategies generally defined as adaptive, non-adaptive and hybrid [19]. Non-adaptive pooling is performed by generating a number of pools according to a predefined combinatorial design, all of the pools are tested in parallel before identifying the positive sample by deconvolution with an algorithm based on the combinatorial design [20–23]. Adaptive strategies are performed in series, data concerning transmission and the results of each test informs which samples will be included in the next test [24–26]. Hybrid methods involve multiple rounds of combinatorial pooling and all samples are tested in parallel during each round [27, 28].

We propose a new method, Hadamard pooling, which is suitable for single round testing when the rate of positivity is low, or which can be used in a hybrid approach when the positivity rate is higher. Hadamard pooling is based upon the orthogonality properties of Hadamard *S* matrices that can identify an individual positive sample in a pool. This method is also compatible with multiplex testing, in which multiple targets can be identified in a single assay, for example by RT-PCR [29, 30], CRISPR-Cas9 assays [31] or colorimetric RT-LAMP analysis [32]. Hadamard pooling has the potential to increase the accuracy and efficiency of surveillance testing programs to track the emergence and spread of infectious diseases in the event of a new pandemic. The idea is explained in fig 1.

**Figure 1.**
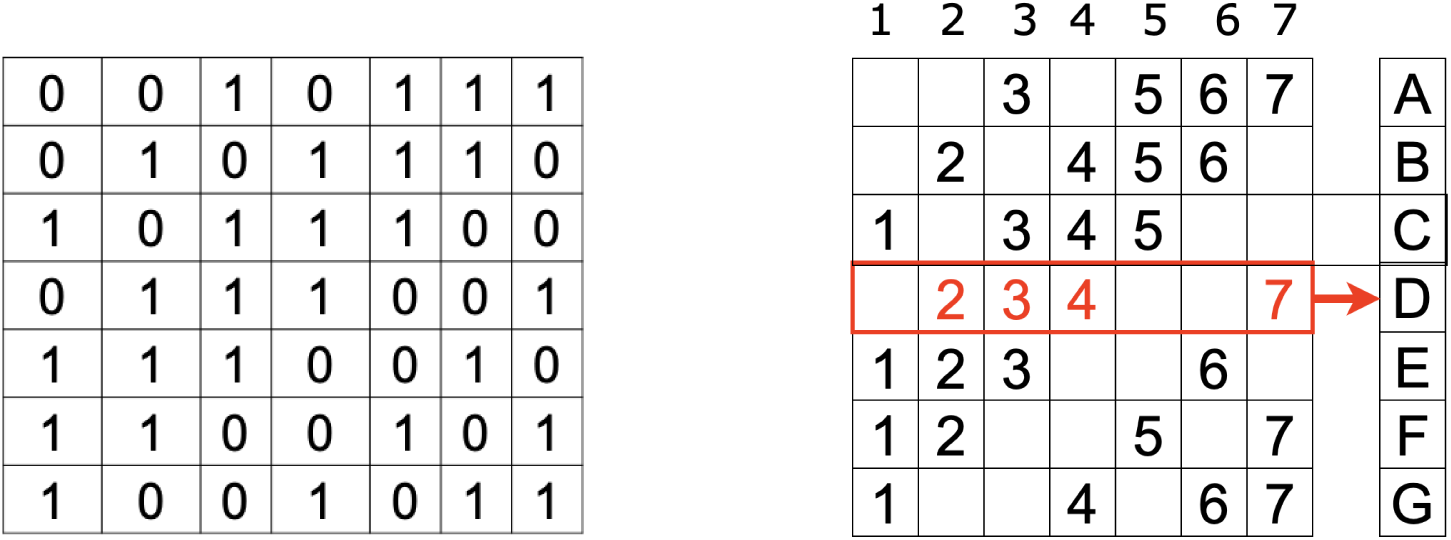
Hadamard pooling strategy scheme based on a 7 × 7 Hadamard matrix (Left). RIght. Pools A - G are generated by grouping together 4 of the 7 individual samples according to the sequence of 1’s and 0’s in each row of the matrix where 1 indicates a sample should be added to the pool and 0 indicates that it is omitted. In the case of pool D (red) samples 2, 3, 4 and 7 would be grouped.

### Hadamard Pooling

Hadamard *S* matrices (Figure 1, Left) have found application in numerous instances of experimental optimal design including signal processing [33–36], imaging [8, 37, 38], X-ray crystallography [39, 40] and spectroscopy [41–46]. In each of these Types of experiments, a signal is modulated by the pattern of a row in the matrix and summed on the detector. After this a linear transform with the inverted Hadamard *S* matrix returns the individual data. By summing up the signal its size is increased, but random noise of detection is averaged towards a constant value and therefore the signal-to-noise can be improved. The process of using the matrices and different ways of generating their patterns is described in the supporting information, S1 appendix, in our online code and in references [39, 40, 43, 47]. Hadamard matrices in general are described in Harwit & Sloan [47].

In the context of pooling methods, individual samples can be grouped into multiple pools according to each row of an *S* matrix with equivalent order to the number of samples being tested. Figure 1 shows a 7 × 7 *S* matrix to illustrate the pooling approach, in practice one of many larger Hadamard matrices could be used, although Hadamard matrices can only be generated with specific sizes, they are numerous.

In the example shown in Figure 1, *n* tests are needed for *n* samples and so there is no improvement over efficiency in comparison to testing individuals. However, in practice only a fraction of the matrix is needed to successfully identify a positive individual. For the purpose of demonstration, a set of seven samples in which one is positive are used. Three pools A, D and F are generated by grouping four of the samples together according to a reduced 3 × 7 matrix constructed from the corresponding three rows of the 7 × 7 *S* matrix (Figure 2).

**Figure 2:**
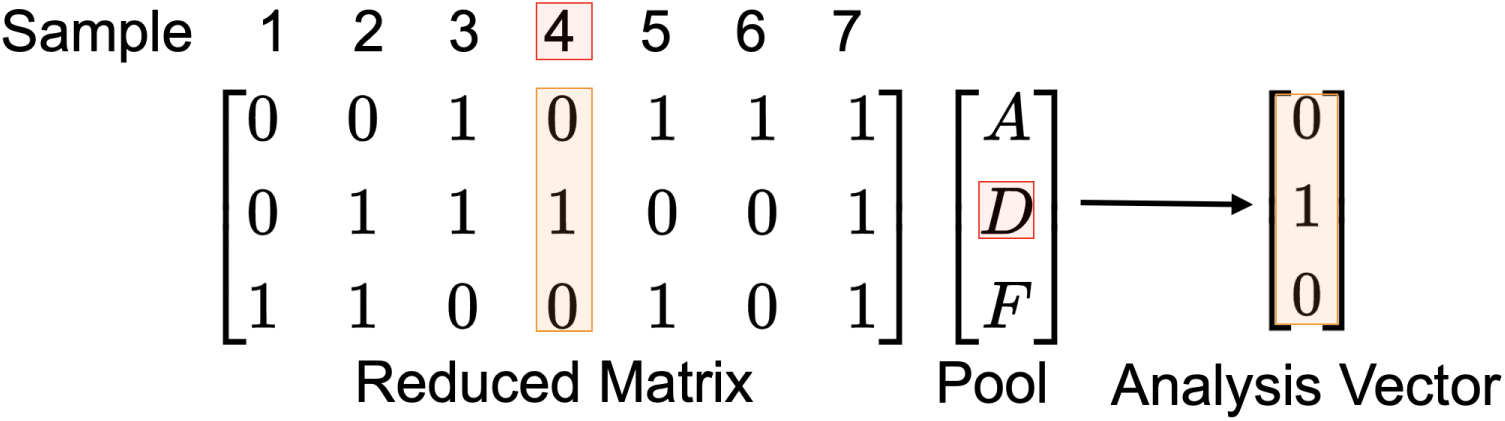
Reduced matrix with samples numbered 1 to 7 grouped into pools A, D and F. Red indicates which sample is positive. The analysis vector elements indicate the results of the pool testing where 1 indicates a positive analysis result and 0 a negative one. Orange highlights the column of the reduced matrix that matches the analysis vector and therefore indicates that sample 4 is positive.

Since the pattern of each column in the matrix is unique it is possible to identify which particular individual is positive and this is done by matching the pattern of results from a specific test to the corresponding column of the reduced matrix. In the example shown in Figure 2, pools A and F return a negative test result, while pool D is positive. The results from each pool are recorded in a vector where 0 indicates a negative pool and 1 indicates a positive pool. This analysis vector is then compared to each column in the reduced matrix, if the analysis vector and column match this indicates that the corresponding sample is positive. For the case shown in Figure 2 the analysis vector, [0 1 0], matches the column in the reduced matrix corresponding to sample 4.

With this method if, for example, eleven specimens need to be checked the nature of the 11 *×* 11 Hadamard matrix is such that only four measurements are needed to confirm a result provided that no more than one specimen is positive. The pattern of the 11 *×* 11 matrix is shown, with others, in the Supporting information. Similarly, assuming that just one sample is positive, only four rows of the *n* = 15 and six of the *n* = 31 Hadamard *S* matrices are needed, Table 1, labelled as Type (i).

**Table 1.** Suitable combinations of Hadamard *S* matrices initially of size *n* x *n*. The last column of the *S* matrix was ignored in choosing lists of rows in Type (ii). In the larger matrices there are equivalent rows that could be used. The right-hand column shows the probability, as a percentage, of detecting 0, 1 or *≥* 2 positives calculated using the Poisson distribution when there are 1% positive samples placed into *n* - 1 bins. This shows the change in percentage of 0, 1, 2 etc. positives in round one of pooling particularly with larger matrices. The letters follow the order ‘A, B, C, . . Z, a, b, c, . . z’.

| $n$ | Row labels.<br>1 positive sample.<br>Type (i) | Row labels,<br>> = 1 positive sample.<br>Type (ii) | Chance (as %) of 0, 1, $\geq 2$<br>present. (Using $n-1$ in the<br>Poisson distribution with<br>probability $p = 0.01$) |
| --- | --- | --- | --- |
| 7 | A D F | A D F G | 94.2, 5.65, 0.173 |
| 11 | B D E F | A C G H I K | 90.5, 9.05, 0.468 |
| 15 | B C D E | A B C D E F I J K O | 86.9, 12.2, 0.893 |
| 19 | B E F H N | A C D I K M N O P S | 83.5, 15.0, 1.44 |
| 23 | A H K N Q | A F H I K O P R T V W | 80.3, 17.7, 2.09 |
| 27 | A B E N S | A B C D E F G H J M O T X | 77.1, 20.1, 2.85 |
| 31 | B C E G L N | C D G J L N P U V X Y a e | 74.1, 22.2, 3.69 |

The condition of low prevalence and hence a low probability of a positive test should be the general expectation when large swathes of a population are tested and the infection is not spreading exponentially i.e. reproduction ratio *R <* 1. The type when more than one sample is positive is examined next as this involves choosing rows more carefully.

### Choosing Rows

If, for example, seven samples are tested using an *S* matrix size of, say 15 × 15, then combinatorially a large number of rows are available to determine which pools to generate, e.g. 15!/(7!(15-7)!) = 6435 and this increases very rapidly with the size of the matrix. Combinations of rows with columns that contain the same pattern of ones and zeros must be excluded. For the example in Figure 1, using pools A, B and D is not suitable because columns 5 and 6 will have the same pattern in the corresponding reduced matrix and therefore the analysis vector cannot distinguish whether sample 5 or 6 is positive.

A further condition is imposed when the number of positive samples is either one or two. Rows must be selected such that when two of their columns are added together, they are not equal to a third. This cannot be achieved if the result is not quantitative, not in an absolute way but that a result containing one positive can be distinguished from that containing two. The consequence of this restriction is that one column of the *S* matrix must be removed (reducing the total number of samples) and in the case of the 7 × 7 matrix one more row is added to the reduced matrix of figure 2, meaning that four pools are now needed from 6 samples. The only acceptable combination of pools for this matrix is A D F G, shown in Figure 3. Table 1 lists rows for other matrices.

**Figure 3:**
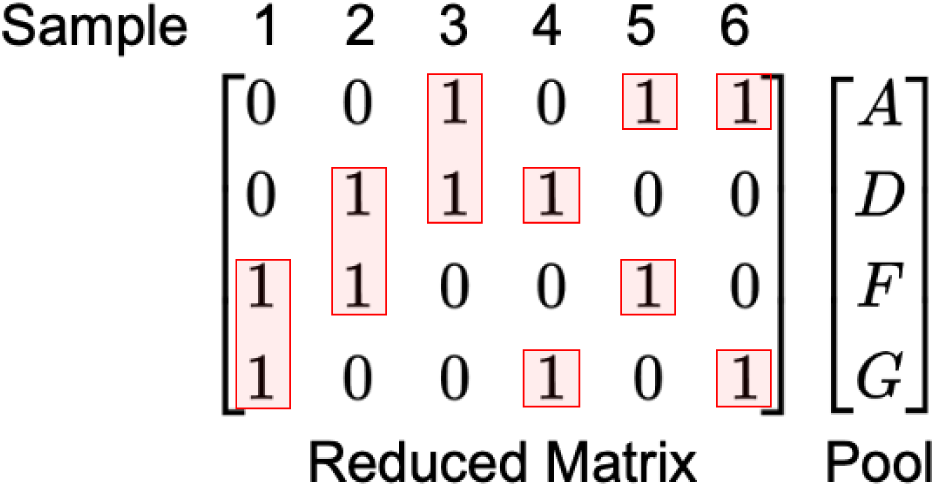
The optimal reduced matrix based on a 7 × 7 Hadamard *S* matrix with corresponding pools. Red shows that no two columns are identical.

There can be several acceptable groups of rows in larger matrices, but still only about 1% of the total combinations. With the 11 x 11 matrix these rows are A C G H I K and in all cases the last column in the Hadamard *S* matrix must be deleted. Different rows will be produced if an alternative column is deleted. Choosing rows so that duplicates are avoided and reducing by one the number of columns and assuming the measurements are relatively quantitative is called Type (ii), see Table 1. Type (i) assumes only one positive is present. When several samples are positive the result will not correspond to any single column in the matrix, in this case all patterns must be checked and these can easily be computed ‘on the fly’ by using a combinatorial function or via a pre-computed look-up table. An example of this is given in table 2 of the SI.

## Methods

### Matrix Generation

All calculations were performed using Python 3 (v 3.9) with the NumPy package (v 1.23) within a Jupyter Notebook (v 6.4). Hadamard matrices were generated using the Quadratic Residue, Shift Register or Doubling methods, as appropriate, and described in [33, 39, 47, 48] and in online code.

To find suitable combinations of the Hadamard matrix rows as shown in Table 1, all the combinations *C_r,n_* = *n*!*/*(*r*!(*n−r*)!) of *r* rows out of a total of *n* were calculated. For example, selecting 10 rows of the *n* = 15 × 15 matrix produces 3003 different combinations, but this rises rapidly to over 200 million for the *n* = 31 matrix with 13 rows. For smaller numbers of combinations each was exhaustively checked. The values of *r* were increased until a satisfactory list was found, see Table 1. There is only one smallest list for a *n* = 11 × 11 matrix, i.e. A C G H I K and 65 smallest but equivalent lists out of 92378 when *r* = 10*, n* = 19 one of which is shown in Table 1. For larger matrices there are also several acceptable lists, but still a small fraction of the total (*<*1%), and only one of them is given in Table 1. (The exception is *n* = 15 where there are many (≈ 13 %) suitable combinations). When the number of combinations runs into millions, as it does for larger matrices, an exhaustive search is not feasible so the combinations were chosen at random until a suitable one was found. This is possible because, luckily, it happens that the suitable combinations tend to cluster and so can be accessed more quickly by random selection than a linear search.

### Pooling Simulation

The test/person ratio was calculated for different sample sizes in two different ways. First using Equation 1 (see below) assuming *m* rows were used out of an *n* x *n* matrix with proportions *f*_0_*, f*_1_ *· · ·* calculated from the Poisson distribution for the percentages of 0, 1, ≥ 2 positives, some of which are given in Table 1. The result is shown in fig 4 as the line with grey circles for 0.1 % positive samples. The second and more detailed simulation started by randomly placing 0.1% or 1% ones into a vector 10000 x *n* long and then splitting this into 10000 groups each of *n* values and pooling these groups according to the two stage Dorfman method. These vectors were then analysed as in an experiment using the matrix rows as listed in Type (ii), Table 1. The resulting tests/person are shown in fig. 4 as the blue circles with 0.1% positive and orange circles with 1% positive. By separately analysing the initial 10000 groups each of length *n* the Binomial nature of the distribution was confirmed for each percentage of positives. These percentages were effectively identical to those produced by the Poisson distribution.

**Figure 4:**
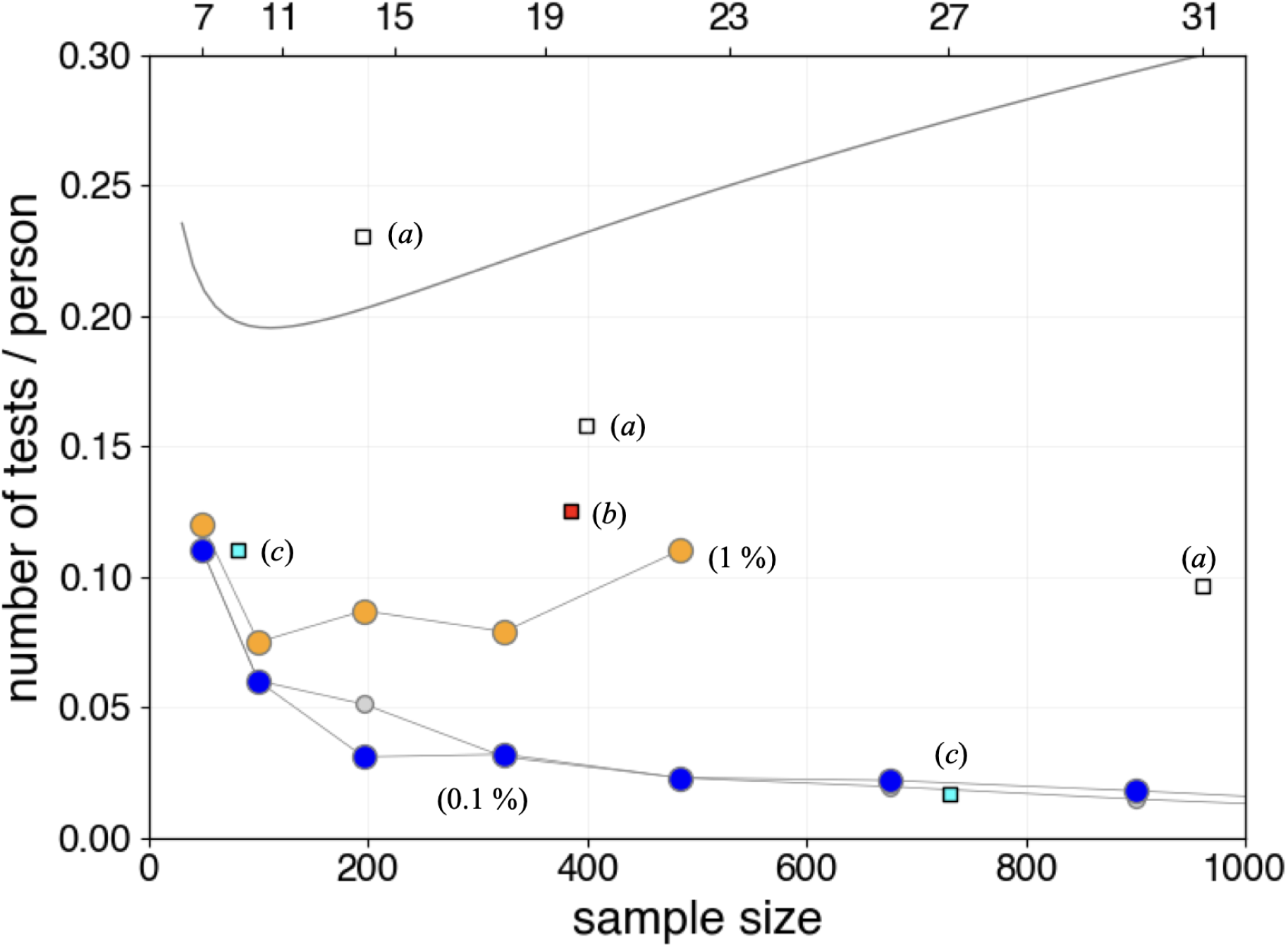
The number of tests / person vs. the sample size (reduced matrix column size squared, (*n* - 1)^2^) in a two step or Dorfman consecutive (two-stage or hierarchical) analysis with 0.1 and 1% positive samples. The top axis shows the *S* matrix size *n*. The blue circles are the full simulated data (see text) when 0.1% on average are positive and the orange circles when 1% are positive. The grey circles at 0.1% positive were calculated using eqn. 1. Points labelled (a) are taken from Ghosh et al. [22], point (b) from Shental et al. [48], and (c) at 1 and 0.1 % positives from Figure 3 in Mutesa et al. [16]. The smooth curve is the Dorfman function.

## Results and Discussion

The Hadamard matrix method is suitable when using Dorfman two-stage pooling and the likelihood of measuring positive samples is low, which here means less than 1%. This limit is the result of the initial pooling greatly increasing the effective percentage of positive samples, thus if the probability is *p* before pooling, when placed into *m* bins it becomes *mp*. In an individual set of data, for example generated using rows of a Hadamard matrix given by Type (ii) of Table 1, if there are two positives then both are correctly identified for each matrix size provided that the samples can be measured quantitatively relative to one another. Finding suitable rows was achieved by exhaustively choosing them and selecting only those that have this property as described in Methods. A consequence of a quantitative measurement is, for example, that if there are 3 positive samples in an initial 11 x 11 matrix all will be detected correctly except that 10 out of the 120 possible ones will need to be fully retested because the pattern produced in these few cases is ambiguous; see Table SI(1) for other examples. In a single measurement this offers only a slight advantage over initially measuring all samples individually, but in the two-stage Dorfman process it is advantageous because, as mentioned, the fraction of positives after pooling is increased.

In Table 1 the first column labelled Type (i) shows the rows that could be used when Dofrman pooling is not used and it is known that exactly one sample is positive and if this is the case then the rows in this column represent the minimum measurements needed to identify this sample. However, in reality this is never the case and there is generally a (Poisson) distribution of positives, 0, 1, 2, etc. In the case when the matrix size chosen is 11, zero positives will be expected in 89.6% of samples, one positive in 9.85% of them and retesting needed for the remaining ≈ 0.56%. The number of tests on average in Type (i) would be approximately 0.995×4 + 0.005×11 ≈ 4, but this is misleading because in Type (i) some results could be ambiguous, i.e. the same pattern can correspond to different positive samples and this will require retesting to sort out. This ambiguity is not present for Type (ii) testing and now ≈ 6 tests are needed but in this case no-retesting is needed for 0, 1, 2 positives and retesting is necessary at only the 0.05% level, (= 0.56 × 10/120, see Table SI(1)) for 3 or more positives. The pattern of rows from Type (ii) is therefore the best option to use.

### Large sample sizes

For a large number of samples with low positivity rate, a strategy similar to that of Dorfman [15] can be used. In this method, all the specimens are pooled into a few large groups and then each of those that tested positive is retested. Our approach is similar but differs slightly in that these initial groups are tested according to the Hadamard method and then the specimens comprising any positive ones are tested again, also using the Hadamard method. Thus for 225 samples (15 x 15 matrix) if only one sample is positive the Dorfman method involves testing 15 pooled groups then 15 more tests of the group containing the positive one. Using the Hadamard method, if only one positive is known to be present and 4 rows, Table 1 Type(i) are used to generate the pools, then four further measurements are needed from the group that contains the positive result, or 2*m* where *m* is the number of rows used. This is 3.6% of the total sample when *n* = 15 or a test/person ratio of 0.036. These values are similar to the 0.1% positives simulation and shown as blue circles in Figure 4.

In the general case the number of positive samples should be present in proportions given by the Poisson distribution. If more than one positive is found in the first round using Type (i), retesting is needed to properly find all positives, which could be by using rows from Type (ii), and then the results from this analysed further if there are any. If *m* rows are initially used from a matrix of size *n* and then *m* rows subsequently and *i* = 0, 1, 2 positive samples occur with the fraction *f_i_* when grouped in the first round and *f*^0^*_i_* in the second round. The number of tests per person *T* when the probability of a positive sample is low is the weighted sum of zero, one, two or three positives etc. and this approximately given by,

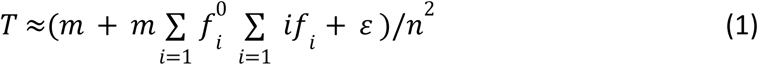

Since the initial test is always made and no further test made when the result is zero, *f*_0_ is absent. A plot of *T* is effectively the same as that from a stochastic simulation when the probability is 0.1%; both are shown in fig. 4. When the percentage of positives is very small indeed, equation 1 is largely given by the first term, so that *T* → *m*/*n*^2^ but when this is not the case the summation becomes important. The term *ɛ* is added and accounts for the fact that at large matrix sizes and at a larger percentage of positive samples, many groupings will contain more than 5 positives, this being approximately the limit of the Hadamard method, see table SI(1). This term is given by 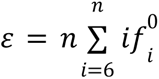, and is only important if the percentage positive is ≥ 1% and the matrix size *n* is large, i.e. when the probability, *f_i_*,(*i* > 5) is no longer very small. The stochastic simulation ratio of tests/person at 1% is also shown in fig 4. The same calculation using eqn. 1 follows this curve.

Data points taken from the literature and the Dorfman function for the number of tests per person are also shown in fig. 4. The Dorfman function is given by equation 2,

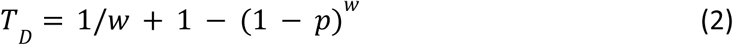

where *w* is the sample size and *p* the probability of infection. The sample size for the minimum number of tests/person is *w*_min_=1/p^1/2^ which is 10 when *p* = 0.01 as seen in fig 4. The number of tests required per person by the Dorfman function subsequently increases with the sample size, in contrast, using the Hadamard method the number of tests decreases for small positivity and is comparable to results of Mutesa et al. [16] for large numbers of samples. At larger positivity, e.g. 1% there is a small decrease with increase in sample size with Hadamard pooling, as with Dorfman pooling, however, the number of tests/person is always significantly lower than that. The Hadamard pooling method produced similar results to previous pooling strategies [16],[28],[49] but has the advantage that no complicated algorithm is needed to interpret the results, only simple pattern matching.

One approach to test the positivity would be to use the minimum number of tests initially (Type (i) Table 1) to find if any of *n* the groups of samples is positive and then use this again for those found to be positive. However, because some ambiguity may arise this will not always identify individuals but instead approximately the number of positives present. With the 11 x 11 matrix and at 1% positivity this would be on average ≈ 4.1 measurements and test/person of 0.04. However, if Type (ii) rows only are used the average number of tests rise to ≈ 6.2 so this may be preferred as all the positives are identified not just their number. An alternative approach could be to test all the *n* samples in round 1, and then test again using Type (i) rows for the second round. This will lead to about twice as many tests per person as using Type(ii) in both rounds (see fig. 4), but could be a good approach if the initial percentage of positives is larger than 1%. Should the full matrix have to be evaluated the simplest way to identify all positive samples using the Hadamard method is to left-multiply the column of results by the inverse of the *S* matrix, see the Supplementary Information.

The pooling method can be extended further for larger numbers, for example with three stages, 15^3^ = 3375 samples, between 12 and 21 measurements are needed. However, this and larger numbers are probably only rarely within the capacity of a single testing station and, although dilution of samples is possible [16, 26], such huge dilution may not be desirable or even feasible in practice.

Finally, we note that the possibility of a false positive or negative can be reduced by carefully choosing the rows, see Table(1). If the data is arranged as a matrix, as in fig. 3 the 4 x 6 matrix (based on the *n* = 7, *S* matrix) only has 2 positives in any column and in a fixed pattern, similarly the 6 × 10 matrix (*n* = 11, *S* matrix) only has 3 values in any column, *n* = 15, has 6 values and *n* = 19, 5 values. A false positive or negative should therefore be detectable.

## Conclusion

We have demonstrated a straightforward approach to pooled testing using Hadamard *S* matrices to design pooling strategies. The efficiency of Hadamard pooling increases with sample size making the approach suitable for surveillance testing in low prevalence settings, ideally when the positivity rate is *≤* 1%. Hadamard pooling not only provides improvements in experimental screening efficiency but also in computational efficiency. A positive sample can be identified from a single round of testing by pattern matching, removing the need for computationally expensive decoding algorithms. A way of testing for false reading is suggested. Our strategy has the potential to contribute to the sustainable and efficient screening programs needed to ensure preparedness for the next pandemic.

## Data Availability

All data produced in the present study are available upon reasonable request to the authors.

## Acknowledgements

We thank Ethan Beddard, Prof. Tia Keyes and Prof. Arwen Pearson for their constructive comments. We also thank Rapid Reviews\Infectious Diseases reviewers for their helpful comments on an earlier version.

## Data and code availability

The code for calculating matrices and rows is included in git commit 2afae1b of the Hadamard repository (https://github.com/byorke/Hadamard).

## Author Contributions

B.A.Y. data analysis, co-wrote the paper. G.S.B. research conception, code, data analysis, co-wrote the paper.

## Competing Interests

The authors declare no competing interests.

## Supporting Information

### Hadamard Matrices

A conventional Hadamard matrix is square and contains entries of 1 and −1 only, and has the property that each row is orthogonal and as such has matching entries in half the rows and half not. The first row and column contain only ones. A variation of these matrices is used here and are labelled as Hadamard **S** matrices in which the first row and column of the Hadamard matrix are ignored and the changes 1 *→* 0 and *−*1 *→* 1 are made in the rest of the matrix. This is the form of the matrix shown in Figure 1.

Only certain integer values are allowed in forming the *S* matrices, nevertheless they are numerous, the first few are

3, 7, 11, 19, 23, 27, 31, 43, 47, 59, · · · 103, · · ·, 199, · · ·

and are prime numbers which also satisfy the condition 4*n* + 3 where *n* = 0, 1, 2*, · · ·*. Other Hadamard matrices can be made with size 2*n −* 1*, n* = 2, 3, for example *n* = 4 produces a 15 *×* 15 matrix and 27 *×* 27 by a similar method. The *n* = 32 and 31 *S* matrices are circulant. Their first rows are:

*n* = 23, 0 0 0 0 1 0 1 0 0 1 1 0 0 1 1 0 1 0 1 1 1 1 1
*n* = 31, 0 0 1 0 0 1 0 0 0 0 1 1 1 0 1 0 1 0 0 0 1 1 1 1 0 1 1 0 1 1 1

### The Hadamard Transform

If the situation arises that all samples have to be measured, a vector, *W*, is constructed out of every summed row in a Hadamard matrix, instead of just selected ones. This vector comprises the unknown values *q_k_* which are multiplied by *S_ik_* = 0 or 1 and summed according to the pattern of each row into:

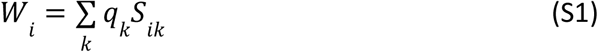

which is the dot product of vectors *q* and a row of *S*. The column vector formed is *W* = [*w*_1_, *w*_2_, …]^T^. The transform to return the original values is

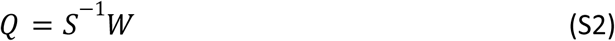

The inverse of the *S* matrix is

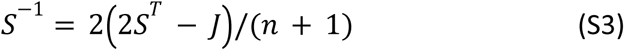

where *J* is the all ones matrix and *T* indicates the transpose.

**Figure S-1.**
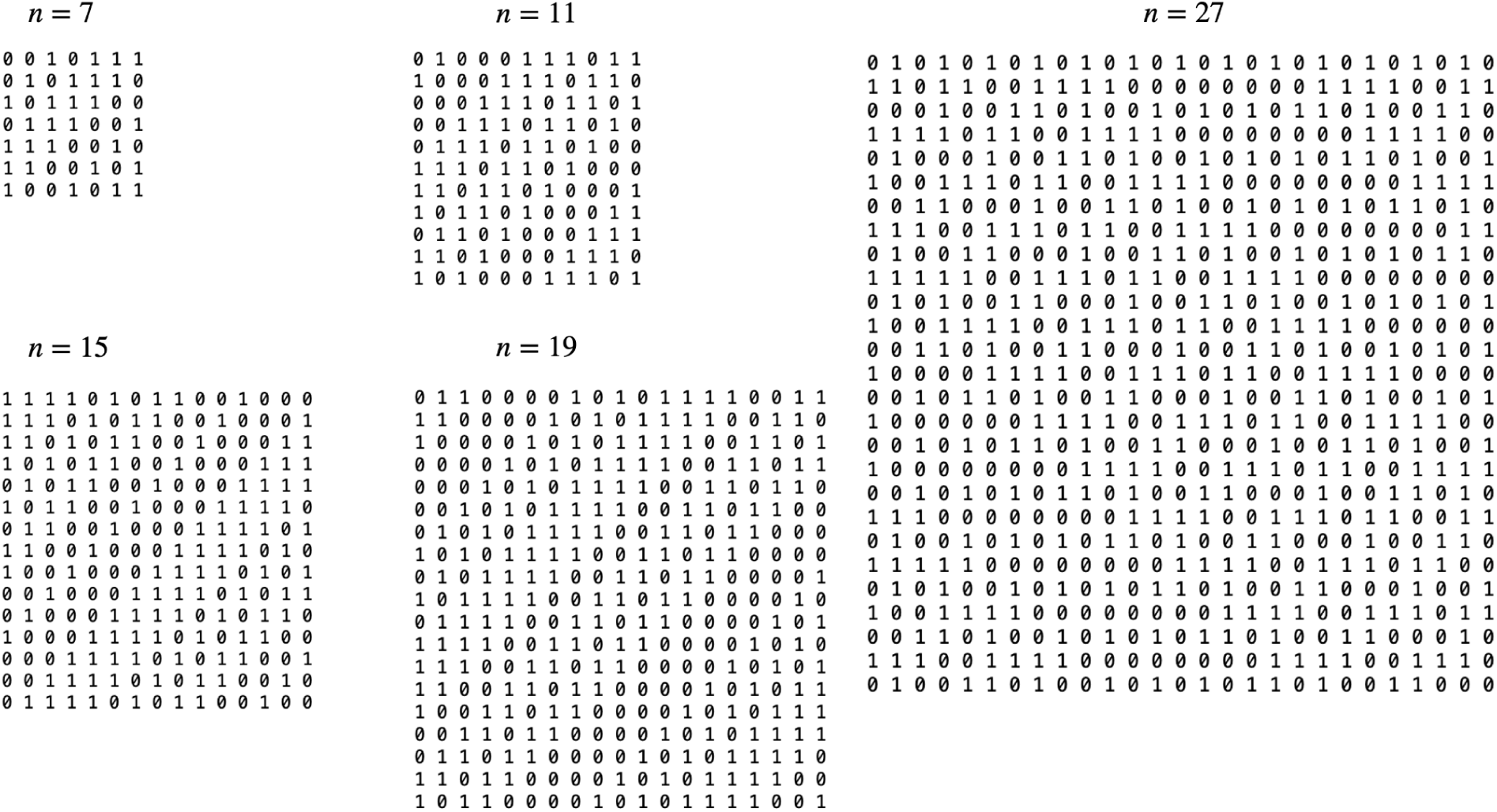
Examples of Hadamard *S* matrices. Each of the matrices except *n* = 28 is circulant and so can be produced from the first lines alone by rotating by one position. Note that the 15 *×* 15 matrix cannot be made by the Quadratic Residue method, however, different but functionally equivalent matrices are formed by either the Shift Register method (matrix shown above) or the Doubling method, both of these methods are described in [46] and the 28 *×* 28 Hadamard matrix construction, among others in refs [33, 47].

**Table SI(1).** The chance or fraction of positive, but duplicate, patterns occurring for each matrix size calculated with Type (ii) rows shown in Table 1. The total number of patterns is calculated as the number of combinations (*C_r,n_*) using *n* - 1 for the matrix size *n*.

| S matrix size →<br>Number positive ↓ | 7 | 11 | 15 | 19 | 23 |
| --- | --- | --- | --- | --- | --- |
| 2 | 1/15 | 0/45 | 0/91 | 0/153 | 0/231 |
| 3 | 6/20 | 10/120 | 0/364 | 0/816 | 0/1540 |
| 4 | 3/15 | 11/210 | 4/1001 | 0/3060 | 0/7315 |
| 5 | 0/6 | 40/252 | 24/2002 | 18/8568 | 0/26334 |
| 6 | - | 65/120 | 57/3003 | 275/18564 | - |

**Table SI(2).**
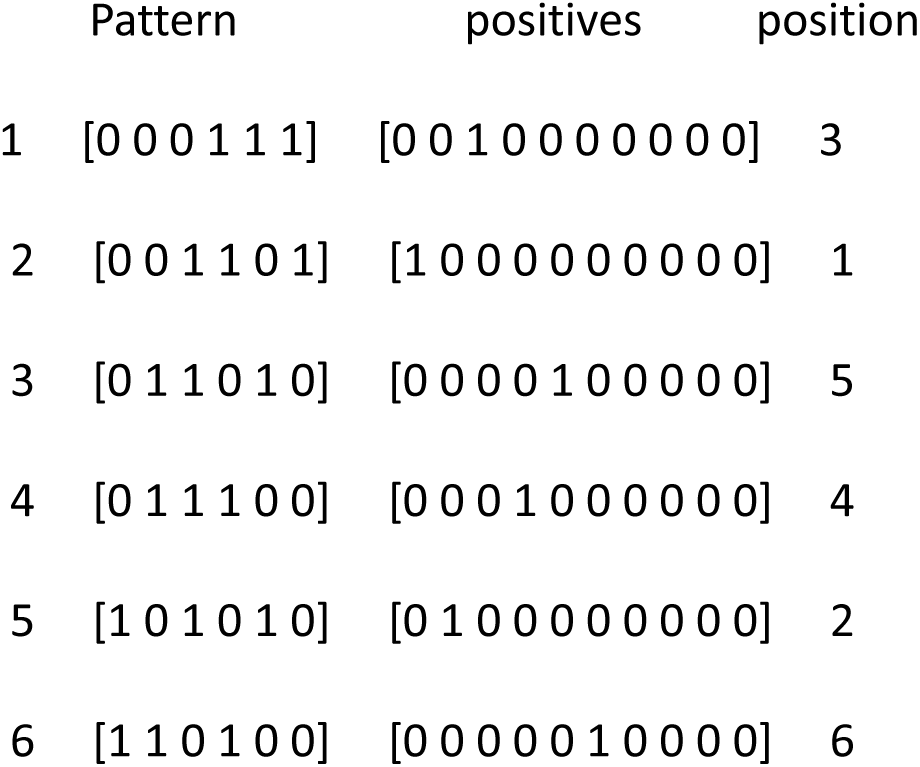
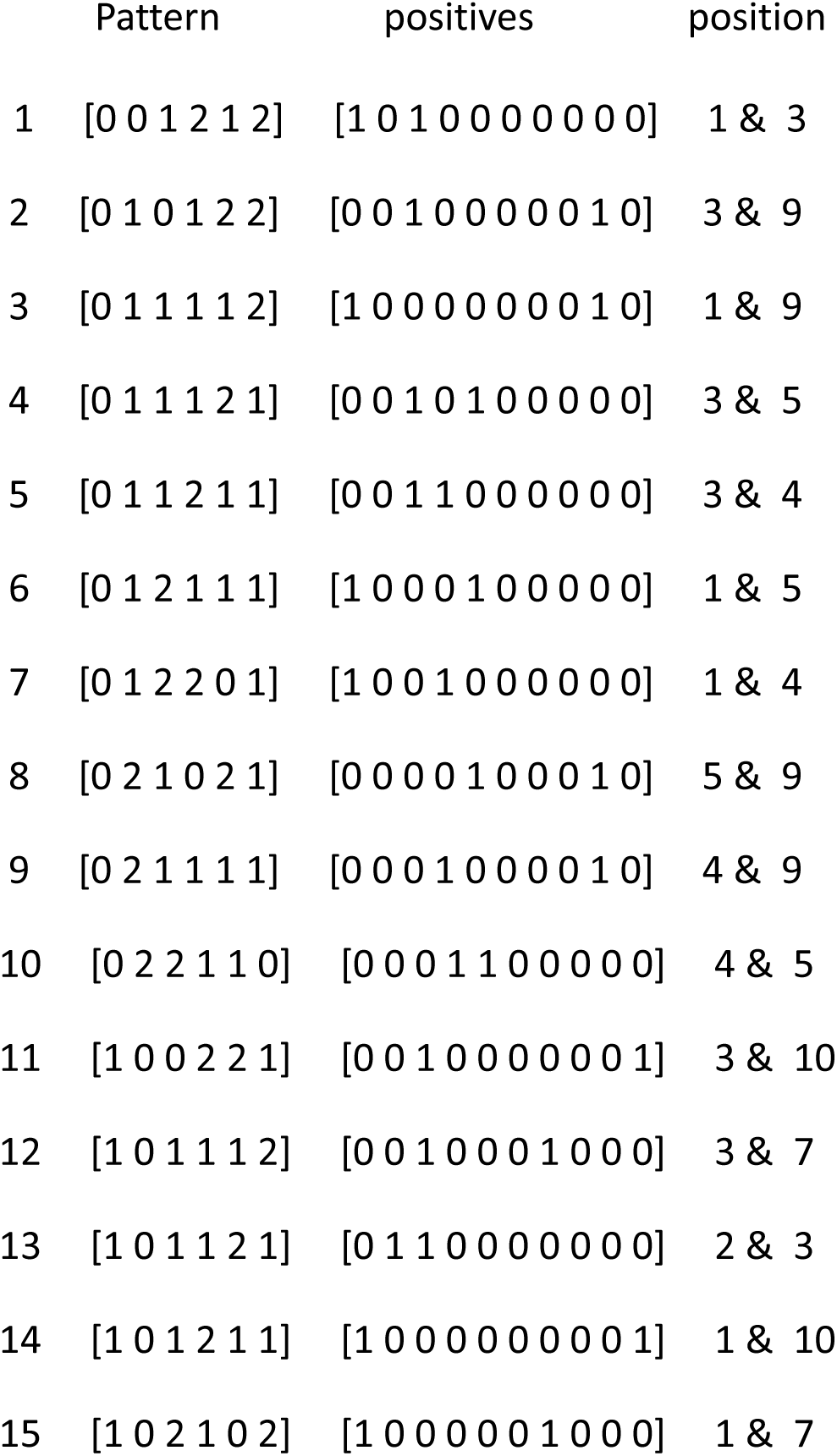
(a) The patterns formed when a 7 matrix is used with 6 columns A D F G, see table 1, and with a single positive sample. The patterns have been sorted and positives positioned accordingly. (b) Example of a look-up table for a 11 x 11 matrix of Type (ii) using the 6 rows A C G H I K, showing part of the list of results, i.e. pattern (vector) observed, and their corresponding position when two positives are present in the ten samples. The patterns have been sorted and positives positioned accordingly.

|  | Pattern | positives | position |
| --- | --- | --- | --- |
| 1 | [0 0 0 1 1 1] | [0 0 1 0 0 0 0 0 0 0] | 3 |
| 2 | [0 0 1 1 0 1] | [1 0 0 0 0 0 0 0 0 0] | 1 |
| 3 | [0 1 1 0 1 0] | [0 0 0 0 1 0 0 0 0 0] | 5 |
| 4 | [0 1 1 1 0 0] | [0 0 0 1 0 0 0 0 0 0] | 4 |
| 5 | [1 0 1 0 1 0] | [0 1 0 0 0 0 0 0 0 0] | 2 |
| 6 | [1 1 0 1 0 0] | [0 0 0 0 0 1 0 0 0 0] | 6 |
| 1 | [0 0 1 2 1 2] | [1 0 1 0 0 0 0 0 0 0] | 1 & 3 |
| 2 | [0 1 0 1 2 2] | [0 0 1 0 0 0 0 0 1 0] | 3 & 9 |
| 3 | [0 1 1 1 1 2] | [1 0 0 0 0 0 0 0 1 0] | 1 & 9 |
| 4 | [0 1 1 1 2 1] | [0 0 1 0 1 0 0 0 0 0] | 3 & 5 |
| 5 | [0 1 1 2 1 1] | [0 0 1 1 0 0 0 0 0 0] | 3 & 4 |
| 6 | [0 1 2 1 1 1] | [1 0 0 0 1 0 0 0 0 0] | 1 & 5 |
| 7 | [0 1 2 2 0 1] | [1 0 0 1 0 0 0 0 0 0] | 1 & 4 |
| 8 | [0 2 1 0 2 1] | [0 0 0 0 1 0 0 0 1 0] | 5 & 9 |
| 9 | [0 2 1 1 1 1] | [0 0 0 1 0 0 0 0 1 0] | 4 & 9 |
| 10 | [0 2 2 1 1 0] | [0 0 0 1 1 0 0 0 0 0] | 4 & 5 |
| 11 | [1 0 0 2 2 1] | [0 0 1 0 0 0 0 0 0 1] | 3 & 10 |
| 12 | [1 0 1 1 1 2] | [0 0 1 0 0 0 1 0 0 0] | 3 & 7 |
| 13 | [1 0 1 1 2 1] | [0 1 1 0 0 0 0 0 0 0] | 2 & 3 |
| 14 | [1 0 1 2 1 1] | [1 0 0 0 0 0 0 0 0 1] | 1 & 10 |
| 15 | [1 0 2 1 0 2] | [1 0 0 0 0 0 1 0 0 0] | 1 & 7 |

## Notes

### Competing Interest Statement

The authors have declared no competing interest.

### Funding Statement

This study did not receive any funding

### Summary of Updates

Updated calculations shown in figure 4. Clarifications given in main text regarding false positives/negatives. Correction of typos. More information provided in supplementary information. Link added to github repository for code.

## References

1. K. Munne, V. Bhanothu, V. Bhor, V. Patel, S. D. Mahale, and S. Pande, ‘Detection of SARS-CoV-2 infection by RT-PCR test: factors influencing interpretation of results’, Virusdisease, vol. 32, no. 2, pp. 187–189, 2021.

2. L. Cheng, L. Lan, M. Ramalingam, J. He, Y. Yang, M. Gao, and Z. Shi, ‘A review of current effective COVID-19 testing methods and quality control’, Archives of Microbiology, vol. 205, no. 6, p. 239, 2023.

3. J. Budd, B. S. Miller, N. E. Weckman, D. Cherkaoui, D. Huang, A. T. Decruz, N. Fongwen, G.-R. Han, M. Broto, C. S. Estcourt, et al., ‘Lateral flow test engineering and lessons learned from COVID-19’, Nature Reviews Bioengineering, vol. 1, no. 1, pp. 13–31, 2023.

4. M. N. Esbin, O. N. Whitney, S. Chong, A. Maurer, X. Darzacq, and R. Tjian, ‘Overcoming the bottleneck to widespread testing: a rapid review of nucleic acid testing approaches for COVID-19 detection’, Rna, vol. 26, no. 7, pp. 771–783, 2020.

5. Y. Peng, P. Wu, A. T. Schartup, and Y. Zhang, ‘Plastic waste release caused by COVID-19 and its fate in the global ocean’, Proceedings of the National Academy of Sciences, vol. 118, no. 47, p. e2111530118, 2021.

6. R. T. Wulff, Y. Qiu, C. Wu, D. P. Calfee, H. K. Singh, I. Hatch, P. A. Steel, J. E. Scofi, L. F. Westblade, and M. M. Cushing, ‘Laboratory interventions to eliminate unnecessary rapid covid-19 testing during a reagent shortage’, American Journal of Clinical Pathology, vol. 158, no. 3, pp. 401–408, 2022.

7. F. Araste, A. D. Bakker, and B. Zandieh-Doulabi, ‘Potential and risks of nanotechnology applications in COVID-19-related strategies for pandemic control’, Journal of Nanoparticle Research, vol. 25, no. 11, p. 229, 2023.

8. C. Hwang, N. Park, E. S. Kim, M. Kim, S. D. Kim, S. Park, N. Y. Kim, and J. H. Kim, ‘Ultra-fast and recyclable dna biosensor for point-of-care detection of SARS-CoV-2 (COVID-19)’, Biosensors and Bioelectronics, vol. 185, p. 113177, 2021.

9. T. Svarc, P. Majeric, D. Feizpour, Jelen, M. Zadravec, T. Gomboc, and R. Rudolf, ‘Recovery study of gold nanoparticle markers from lateral flow immunoassays’, Materials, vol. 16, p. 5770, 2023.

10. A. Sun, P. Voparilova, X. Liu, et al., ‘An integrated microfluidic platform for nucleic acid testing’, Microsyst. Nanoeng., vol. 10, p. 66, 2024.

11. W. Cui, P. Zhao, J. Wang, N. Qin, E. A. Ho, and C. L. Ren, ‘Reagent free detection of SARS-CoV-2 using an antibody-based microwave sensor in a microfluidic platform’, Lab on a Chip, vol. 22, no. 12, p. 2307–2314, 2022.

12. M. R. Jamiruddin, B. A. Meghla, D. Z. Islam, T. A. Tisha, S. S. Khandker, M. U. Khondoker, M. A. Haq, N. Adnan, and M. Haque, ‘Microfluidics technology in SARS-CoV-2 diagnosis and beyond: A systematic review’, Life (Basel), vol. 12, no. 5, p. 649, 2022.

13. H. Yousefi, A. Mahmud, D. Chang, J. Das, S. Gomis, J. B. Chen, H. Wang, T. Been, L. Yip, E. Coomes, et al., ‘Detection of SARS-CoV-2 viral particles using direct, reagent-free electrochemical sensing’, Journal of the American Chemical Society, vol. 143, no. 4, pp. 1722–1727, 2021.

14. K. Ember, F. Daoust, M. Mahfoud, F. Dallaire, E. Z. Ahmad, T. Tran, A. Plante, M.-K. Diop, T. Nguyen, A. St-Georges-Robillard, et al., ‘Saliva-based detection of COVID-19 infection in a real-world setting using reagent-free raman spectroscopy and machine learning’, Journal of Biomedical Optics, vol. 27, no. 2, pp. 025002, 2022.

15. R. Dorfman, ‘The detection of defective members of large populations’, Annals of Mathematical Statistics, vol. 14, p. 436, 1943.

16. L. Mutesa, P. Ndishimye, Y. Butera, J. Souopgui, A. Uwineza, R. Rutayisire, E. L. Ndoricimpaye, E. Musoni, N. Rujeni, T. Nyatanyi, D. Nyamwasa, M. Ndashimye, E. Ujeneza, I. E. Mwikarago, C. M. Muvunyi, J. B. Mazarati, S. Nsanzimana, N. Turok, W. Ndifon, ‘A pooled testing strategy for identifying SARS-CoV-2 at low prevalence’, Nature, vol. 589, no. 7841, pp. 276–280, 2021.

17. M. Aldridge and D. Ellis, ‘Pooled testing and its applications in the COVID-19 pandemic’, Pandemics: Insurance and Social Protection, pp. 217–249, 2022.

18. J. Burtniak, A. Hedley, K. Dust, P. Van Caeseele, J. Bullard, and D. R. Stein, ‘Dorfman pooling enhances SARS-CoV-2 large-scale community testing efficiency’, PLOS Global Public Health, vol. 3, no. 4, p. e0001793, 2023.

19. T. N. Furstenau, J. H. Cocking, C. M. Hepp, and V. Y. Fofanov, ‘Sample pooling methods for efficient pathogen screening: Practical implications’, PLOS One, vol. 15, no. 11, p. e0236849, 2020.

20. J. Yi, R. Mudumbai, and W. Xu, ‘Low-cost and high-throughput testing of COVID-19 viruses and antibodies via compressed sensing: System concepts and computational experiments’, arXiv preprint arXiv:2004.05759, 2020.

21. N. Shental, S. Levy, V. Wuvshet, S. Skorniakov, B. Shalem, A. Ottolenghi, Y. Greenshpan, R. Steinberg, A. Edri, R. Gillis, et al., ‘Efficient high-throughput SARS-CoV-2 testing to detect asymptomatic carriers’, ScienceAdvances, vol. 6, no. 37, p. eabc5961, 2020.

22. S. Ghosh, A. Rajwade, S. Krishna, N. Gopalkrishnan, T. Schaus, A. Chakravarthy, S. Varahan, V. Appu, R. Ramakrishnan, S. Ch, M. Jindal, V. Bhupathi, A. Gupta, A. Jain, R. Agarwal, S. Pathak, M. Rehan, S. Consul, Y. Gupta, N. Gupta, P. Agarwal, R. Goyal, V. Sagar, U. Ramakrishnan, S. Krishna, P. Yin, D. Palakodeti, and M. Gopalkrishnan, ‘Tapestry: A single-round smart pooling technique for COVID-19 testing’, medRxiv, 2020.

23. J. H. McDermott, D. Stoddard, P. J. Woolf, J. M. Ellingford, D. Gokhale, A. Taylor, L. A. Demain, W. G. Newman, and G. Black, ‘A nonadaptive combinatorial group testing strategy to facilitate health care worker screening during the severe acute respiratory syndrome coronavirus-2 (SARS-CoV-2) outbreak’, The Journal of Molecular Diagnostics, vol. 23, no. 5, pp. 532–540, 2021.

24. M. Escobar, G. Jeanneret, L. Bravo-Sanchez, A. Castillo, C. Gomez, D. Valderrama, M. Roa, J. Martınez, J. Madrid-Wolff, M. Cepeda, M. Guevara-Suarez, O. L. Sarmiento, A. Medaglia, M. Forero-Shelton, M. Velasco, J. M. Pedraza, R. Laajaj, S. Restrepo, and P. Arbelaez, ‘Smart pooling: AI-powered COVID-19 informative group testing’, Scientific Reports, vol. 12, no. 1, p. 6519, 2022.

25. J. Zhang and L. S. Heath, ‘Adaptive group testing strategy for infectious diseases using social contact graph partitions’, Scientific Reports, vol. 13, no. 1, p. 12102, 2023.

26. N. Barak, R. Ben-Ami, T. Sido, A. Perri, A. Shtoyer, M. Rivkin, T. Licht, A. Peretz, J. Magenheim, I. Fogel, et al., ‘Lessons from applied large-scale pooling of 133,816 SARS-CoV-2 RT-PCR tests’, Science Translational Medicine, vol. 13, no. 589, p. eabf2823, 2021.

27. N. Lagopati, P. Tsioli, I. Mourkioti, A. Polyzou, A. Papaspyropoulos, A. Zafiropoulos, K. Evangelou, G. Sourvinos, and V. G. Gorgoulis, ‘Sample pooling strategies for SARS-CoV-2 detection’, Journal of virological methods, vol. 289, p. 114044, 2021.

28. A. Chakravarthy, S. Krishna, S. Ghosh, A. Tomar, S. Varahan, A. Rajwade, S. Ghosh, N. Gupta, R. Agarwal, H. Payal, P. Chakraborty, K. V. Vemula, A. Vyas, R. Goru, S. Krishna, D. Palakodeti, and M. Gopalkrishnan, ‘Large-scale testing for SARS-CoV-2 using tapestry pooling’, medRxiv, 2020.

29. I. A. Correa, T. de Souza Rodrigues, A. Queiroz, L. Nascimento, T. Wolff, R. Akamine, S. Kuriyama, L. da Costa, and A. Fidalogo-Neto, ‘Boosting SARS-CoV-2 detection combining pooling and multiplex strategies’, Scientific Reports, vol. 12, p. 8684, 2022.

30. K. S. Butler, B. D. Carson, J. D. Podlevsky, C. M. Mayes, J. M. Rowland, D. Campbell, J. B. Ricken, G. Wudiri, and J. A. Timlin, ‘Singleplex, multiplex and pooled sample real-time RT-PCR assays for detection of SARS-CoV-2 in an occupational medicine setting’, Scientific Reports, vol. 12, no. 1, p. 17733, 2022.

31. N. L. Welch, M. Zhu, C. Hua, J. Weller, M. E. Mirhashemi, T. G. Nguyen, S. Mantena, M. R. Bauer, B. M. Shaw, C. M. Ackerman, et al., ‘Multiplexed crispr-based microfluidic platform for clinical testing of respiratory viruses and identification of SARS-CoV-2 variants’, Nature Medicine, vol. 28, no. 5, pp. 1083–1094, 2022.

32. D. Lee, C.-H. Chu, and A. F. Sarioglu, ‘Point-of-care toolkit for multiplex molecular diagnosis of SARS-CoV-2 and influenza a and b viruses’, Acs Sensors, vol. 6, no. 9, pp. 3204–3213, 2021.

33. K. J. Horadam, Hadamard matrices and their applications. Princeton University Press, 2012.

34. L. Ou, S. Liao, Z. Qin, and H. Yin, ‘Millimeter wave wireless Hadamard image transmission for mimo-enabled 5g and beyond’, IEEE Wireless Communications, vol. 27, no. 6, pp. 134–139, 2020.

35. A. Aung, B. P. Ng, and S. Rahardja, ‘Sequency - ordered complex Hadamard transform: Properties, computational complexity and applications’, IEEE Transactions on Signal processing, vol. 56, no. 8, pp. 3562–3571, 2008.

36. D. e. Jabeen, G. Monir, and F. Azim, ‘Sequency domain signal processing using complex Hadamard transform’, Circuits, Systems, and Signal Processing, vol. 35, pp. 1783–1793, 2016.

37. C. Oliver and E. Pike, ‘Multiplex advantage in the detection of optical images in the photon noise limit’, Applied Optics, vol. 13, no. 1, pp. 158–161, 1974.

38. Z. Zhang, X. Wang, G. Zheng, and J. Zhong, ‘Hadamard single-pixel imaging versus Fourier single-pixel imaging’, Optics Express, vol. 25, no. 16, pp. 19619–19639, 2017.

39. B. A. Yorke, G. S. Beddard, R. L. Owen, and A. R. Pearson, ‘Time-resolved crystallography using the Hadamard transform’, Nature methods, vol. 11, no. 11, pp. 1131–1134, 2014.

40. M.A. Klureza, Y. Pulnova, D. von Stetten, R.L. Owen, G.S. Beddard, A.R. Pearson, B.A. Yorke, ‘Multiplexing methods in dynamic protein crystallography’ Methods Enzymology, vol. 709, pp. 177–206, 2024.

41. P. Jacquinot, ‘The luminosity of spectrometers with prisms, gratings, or Fabry-Perot etalons’, JOSA, vol. 44, no. 10, pp. 761–765, 1954.

42. J. F. James and R. S. Sternberg, ‘The Design of Optical Spectrometers’, Publ. Chapman and Hall, 1969.

43. G. S. Beddard and B. A. Yorke, ‘Pump–probe spectroscopy using the Hadamard transform’, Applied Spectroscopy, vol. 70, no. 8, pp. 1292–1299, 2016.

44. Y. Zhang, M. O. Malik, J. Kang, C. Yuen, and Q. Liu, ‘Sequency encoding single pixel spectroscopy based on Hadamard transform’, Optics Express, vol. 30, no. 17, pp. 30121–30134, 2022.

45. V. Yatsyna, A. H. Abikhodr, A. Ben Faleh, S. Warnke, and T. R. Rizzo, ‘High-throughput multiplexed infrared spectroscopy of ion mobility-separated species using Hadamard transform’, Analytical Chemistry, vol. 94, no. 6, pp. 2912–2917, 2022.

46. Q. Xu, N. Ding, D. Ma, H. Lin, B. Lin, X. Ma, J. Yang, and L. Guo, ‘Portable Hadamard transform Raman spectrometer: A powerful analytical tool for point-of-care testing’, Analytical Chemistry, vol. 96, no. 30, pp. 12217–12224, 2024.

47. M. Harwit and N. J. Sloane, Hadamard Transform Optics. Academic Press, 1979.

48. A. Hedayat and W. D. Wallis, ‘Hadamard matrices and their applications’, The Annals of Statistics, pp. 1184–1238, 1978.

49. H. Shani-Narkiss, O. D. Gilday, N. Yayon, and I. D. Landau, ‘Efficient and practical sample pooling for high-throughput pcr diagnosis of COVID-19’ MedRxiv, pp. 2020–04, 2020.

